# COVID-19 Vaccine Effectiveness by Product and Timing in New York State

**DOI:** 10.1101/2021.10.08.21264595

**Authors:** Eli S. Rosenberg, Vajeera Dorabawila, Delia Easton, Ursula E. Bauer, Jessica Kumar, Rebecca Hoen, Dina Hoefer, Meng Wu, Emily Lutterloh, Mary Beth Conroy, Danielle Greene, Howard A. Zucker

## Abstract

**Background:** US population-based data on COVID-19 vaccine effectiveness (VE) for the 3 currently FDA-authorized products is limited. Whether declines in VE are due to waning immunity, the Delta variant, or other causes, is debated.

**Methods:** We conducted a prospective study of 8,834,604 New York adults, comparing vaccine cohorts defined by product, age, and month of full-vaccination to age-specific unvaccinated cohorts, by linking statewide testing, hospital, and vaccine registry databases. VE was estimated from May 1, 2021 for incident laboratory-confirmed COVID-19 cases (weekly life-table hazard rates through September 3) and hospitalizations (monthly incidence rates through August 31).

**Results:** 155,092 COVID-19 cases and 14,862 hospitalizations occurred. Estimated VE for cases declined contemporaneously across age, products, and time-cohorts, from high levels beginning May 1 (1.8% Delta variant prevalence), to a nadir around July 10 (85.3% Delta), with limited changes thereafter (>95% Delta). Decreases were greatest for Pfizer-BioNTech (−24.6%, −19.1%, −14.1% for 18-49, 50-64 years, and ≥65 years, respectively), and similar for Moderna (−18.0%, −11.6%, −9.0%, respectively) and Janssen (−19.2%, −10.8, −10.9%, respectively). VE for hospitalization for adults 18-64 years was >86% across cohorts, without time trend. Among persons ≥65 years, VE declined from May to August for Pfizer-BioNTech (95.0% to 89.2%) and Moderna (97.2% to 94.1%). VE was lower for Janssen, without trend, ranging 85.5%-82.8%.

**Conclusions:** Declines in VE for cases may have been primarily driven by factors other than waning. VE for hospitalizations remained high, with modest declines limited to Pfizer-BioNTech and Moderna recipients ≥65 years, supporting targeted booster dosing recommendations.

## Introduction

In New York State (NYS), over 2.4 million people have been diagnosed, and over 56,000 have died, with COVID-19 through Sept 29, 2021.^1^ COVID-19 vaccines are a critical prevention tool, with the three currently FDA-approved or authorized products originally shown in trials to be highly-efficacious against moderate-severe disease among adults, with mRNA-BNT162b2 (Pfizer-BioNTech) at 95%, mRNA-1273 (Moderna) at 94%, and Ad26.COV2 (Janssen) at 72% efficacy.^2-4^ These studies’ endpoints were evaluated at relatively short follow-up of 14-28 days after series completion and during a period when circulating strains were less transmissible than the currently-predominant Delta variant.^5^

More recently, extended trial follow-up and real-world effectiveness studies have begun to document declines in efficacy and vaccine effectiveness (VE), for some outcomes and populations, during 2021. Studies in Israel documented larger declines in VE than observed in the U.S. for infection and severe disease among its Pfizer-receiving population, which may be due to earlier vaccination, increased sensitivity definitions, or other methodological differences.^6-9^ U.S. population-based effectiveness studies in NYS and sentinel populations have documented declines in VE, particularly for persons ≥65 years, during the period in which the Delta variant became predominant and mitigation strategies and policies were relaxed.^10-14^ The limitations of open cohort surveillance studies and limited numbers of events in more controlled studies make it difficult to isolate and quantify the extent to which VE declines can be attributed to immunologic waning, Delta variant, behavioral changes, or other causes, particularly across subgroups defined by age, product, and risks for infection or severe illness.

Data are urgently needed to understand the magnitude and sources of changes in VE, across outcomes, products, and population subgroups, to inform public health policy and vaccine recommendations. Following U.S. Food and Drug Administration (FDA) authorization, the Centers for Disease Control and Prevention (CDC) recently recommended Pfizer-BioNTech booster doses for persons ≥65 years, those with underlying conditions between 18-64 years, and persons 18-64 years in high-exposure occupations or settings.^15,16^ It is anticipated that additional eligibility expansion and booster doses for Moderna and Janssen will be considered in Fall 2021. In the September 2021 advisory meetings for FDA and CDC, gaps in U.S. VE data by age, time, and vaccine product were notable and a point of debate.^17,18^

To address these gaps, we conducted a statewide cohort study to describe VE in NYS, in closed cohorts defined by product, age, and time of vaccination, from the time of the Delta variant’s emergence to predominance.

## Methods

We conducted a surveillance-based prospective cohort study of NYS adults, by linking statewide immunization, laboratory testing, and hospitalization databases. Incident laboratory-confirmed COVID diagnosis and hospitalization were ascertained among closed cohorts of vaccine recipients, defined by age, product, and timing of full-vaccination, during the pre- vs. post-Delta variant period from May 1 (<2%) to August 28, 2021 (>99%), and compared to the cohort of persons remaining unvaccinated.

### Data sources

Four databases were linked to construct a surveillance-based cohort of adults aged ≥18 years residing in NYS.^10^ The Citywide Immunization Registry (CIR) and the NYS Immunization Information System (NYSIIS) are used to collect and store all COVID-19 provider vaccination data for persons residing in New York City and the rest of the state, respectively (excluding settings that report directly to the federal system such as veterans, military, and first nations tribal healthcare facilities). The Electronic Clinical Laboratory Reporting System (ECLRS) collects all reportable COVID-19 test results (nucleic acid amplification test [NAAT] or antigen) in NYS.^19^ The Health Electronic Response Data System (HERDS) includes a statewide, daily electronic survey of all inpatient facilities in NYS, which collects all new admissions with a laboratory-confirmed COVID-19 diagnosis.

The NYSIIS/CIR COVID-19 data were combined and deduplicated based on first name, last name, date of birth (DOB), and ZIP code then matched to ECLRS using a deterministic algorithm based on first name, last name, and DOB, and to HERDS based on initials, sex, DOB, and residence ZIP code.

### Cohort construction

The first NYS-administered dose for Pfizer-BioNTech was on December 14, 2020, Moderna on December 18, 2020, Janssen on March 4, 2021. Starting December 2020, vaccine eligibility sequentially expanded to priority groups, including those in long-term care facilities, healthcare workers, ≥65 years, frontline and essential workers, and school staff, and those with co-morbidities (Table S1). Closed cohorts were defined using persons fully-vaccinated as of May 1, 2021, on the basis of age (18-49, 50-64, and ≥65 years) and vaccine product (Pfizer-BioNTech, Moderna, Janssen). Cohorts were defined based on timing of full-vaccination (≥14 days after receipt of the final dose): those fully-vaccinated in January-February (Pfizer-BioNTech and Moderna only), March, and April 2021.^20^ Whereas vaccinated cohorts were directly observed in NYSIIS/CIR, three synthetic age-specific unvaccinated comparison cohorts were defined as *Census population minus persons partially or fully vaccinated on September 23*, with case and hospitalization outcomes classified as unvaccinated if no matching COVID-19 vaccine was found in NYSIIS/CIR.

### Statistical approach

For the person-level cases outcome, the time to first new positive SARS-CoV-2 NAAT or antigen test result (based on collection date) was assessed using the life-table method, with 7-day time intervals, from the May 1 to August 28 weeks (ending September 3). For each product and age group, weekly hazard rates and 95% confidence intervals (CI) were estimated overall and by time cohort, with hazard ratios and 95% CI compared to the corresponding age-specific unvaccinated cohort.^21^ VE was estimated as *1-HR*.

The hospitalization outcome was defined as a new admission with a laboratory-confirmed positive COVID-19 result, between May and August 2021. Because the unit of observation was an admission rather than person (approximately 9% of admissions estimated non-unique on persons), an aggregate rates approach was used, as done elsewhere.^10,11^ For each product and age group, the incidence rates of new admissions per population, and exact Poisson 95% CI, were estimated overall by time cohort monthly, given data sparseness at the week-level. Incidence rate-ratios (IRR) and exact binomial 95% CI compared each vaccinated cohort to the respective age-specific unvaccinated cohort, with VE estimated as *1-IRR*.

Trends in VE for cases were graphically compared to the percentage of specimens with the Delta variant from the CDC national SARS-CoV-2 genomic surveillance program, for the HHS region containing NYS (data extracted 9/14/2021).^5^ The association between VE and the percentage Delta variant was summarized using the Pearson correlation coefficient (*r*) at each week, after each was linearized using the logit transformation.

### Primary and sensitivity analyses

In the primary analysis, persons with a positive laboratory result within 90 days before May 1 were considered not susceptible for either outcome and excluded, per the CDC case-definition. Sensitivity analyses probed the impact of variations in the cohort definitions (Supplementary Methods).

## Results

Cohorts included in the analysis, and their outcomes, are summarized in Table 1. Among a total of 8,834,604 adults in this analysis, 5,787,817 (65.5%) were fully-vaccinated. Among those fully-vaccinated, 48.6% received Pfizer, 41.5% Moderna, and 10.0% Janssen. During follow-up, fully-vaccinated persons experienced a total of 38,778 cases and 2,363 hospitalizations and unvaccinated persons had 116,314 cases and 38,778 hospitalizations.

**Table 1.** Total persons, and total new laboratory-confirmed COVID-19 cases and hospitalizations during follow-up, among cohorts of fully-vaccinated and unvaccinated adults in New York State.

| *Vaccine cohort* | Persons | Cases | Hospitalizations |
| --- | --- | --- | --- |
|  |  | *Weeks of May 1 – August 28,*<br>*2021* | *May - August* |
| **<u>18-49 years</u>** |  |  |  |
| **Pfizer-BioNTech** | **980,353** | **10,738** | **95** |
| January/February | 233,998 | 2,991 | 24 |
| March | 157,885 | 1,842 | 12 |
| April | 588,470 | 5,905 | 59 |
| **Moderna** | **752,322** | **5,658** | **60** |
| January/February | 254,209 | 2,206 | 14 |
| March | 180,876 | 1,444 | 15 |
| April | 317,237 | 2,008 | 31 |
| **Janssen** | **276,481** | **3,307** | **38** |
| March | 52,223 | 826 | 14 |
| April | 224,258 | 2,481 | 24 |
| **Unvaccinated** | **2,070,251** | **85,667** | **4,689** |
| **<u>50-64 years</u>** |  |  |  |
| **Pfizer-BioNTech** | **846,664** | **5,602** | **228** |
| January/February | 139,146 | 1,133 | 44 |
| March | 139,140 | 1,076 | 40 |
| April | 568,378 | 3,393 | 144 |
| **Moderna** | **624,226** | **2,723** | **104** |
| January/February | 153,965 | 909 | 19 |
| March | 124,454 | 663 | 10 |
| April | 345,807 | 1,151 | 75 |
| **Janssen** | **184,120** | **1,406** | **92** |

| **Vaccine cohort** | **Persons** | **Cases**<br>*Weeks of May 1 – August 28,*<br>*2021* | **Hospitalizations**<br>*May - August* |
| --- | --- | --- | --- |
| March | 55,292 | 491 | 27 |
| April | 128,828 | 915 | 65 |
| **Unvaccinated** | **606,411** | **20,175** | **3,380** |
| **<u>≥65 years</u>** |  |  |  |
| **Pfizer-BioNTech** | **984,464** | **5,302** | **972** |
| January/February | 202,305 | 1,423 | 249 |
| March | 350,022 | 1,972 | 329 |
| April | 432,137 | 1,907 | 394 |
| **Moderna** | **1,023,748** | **3,291** | **545** |
| January/February | 145,042 | 649 | 90 |
| March | 435,849 | 1,448 | 216 |
| April | 442,857 | 1,194 | 239 |
| **Janssen** | **115,439** | **751** | **229** |
| March | 49,515 | 344 | 88 |
| April | 65,924 | 407 | 141 |
| **Unvaccinated** | **370,125** | **10,472** | **4,430** |
| **Total** | **8,834,604** | **155,092** | **14,862** |

Incident cases declined for all cohorts defined by age, vaccination status, vaccine product and time cohorts from May 1 through late June and increased thereafter following increases in the prevalence of the Delta variant (Figure S1), with rates consistently highest for the unvaccinated. Across product and time cohorts, estimated VE generally declined from highest levels during the week of May 1 (1.8% Delta variant) to a nadir around July 10 (85.3% Delta variant), with modest changes in following weeks (>95% Delta variant) (Table 2, Figure 1).

**Table 2:** Estimated vaccine effectiveness for laboratory-confirmed COVID-19 cases.

| ***Vaccine cohort*** | **May 1 week**<br>**% VE (95% CI)** | **July 10 week**<br>**% VE (95% CI)** | **August 28 week**<br>**% VE (95% CI)** |
| --- | --- | --- | --- |
| **<u>18-49 years</u>** |  |  |  |
| **Pfizer-BioNTech** | **93.6 (92.6, 94.6)** | **65.8 (62.2, 69.5)** | **69.0 (67.4, 70.6)** |
| January/February | 90.5 (88.0, 93.0) | 64.2 (56.9, 71.5) | 67.4 (64.2, 70.6) |
| March | 90.8 (87.9, 93.8) | 64.7 (55.9, 73.4) | 66.2 (62.3, 70.1) |
| April | 95.6 (94.5, 96.7) | 66.8 (62.3, 71.4) | 70.4 (68.5, 72.4) |
| **Moderna** | **96.5 (95.6, 97.3)** | **77.2 (73.9, 80.5)** | **78.4 (77.0, 79.9)** |
| January/February | 94.0 (92.1, 95.9) | 79.8 (74.5, 85.0) | 72.1 (69.3, 74.9) |
| March | 97.8 (96.5, 99.2) | 74.9 (68.1, 81.8) | 79.2 (76.3, 82.0) |
| April | 97.7 (96.6, 98.7) | 76.4 (71.3, 81.5) | 83.1 (81.2, 85.1) |
| **Janssen** | **89.4 (87.0, 91.8)** | **51.7 (43.8, 59.6)** | **70.2 (67.4, 73.0)** |
| March | 90.3 (85.0, 95.6) | 19.8 (0.0, 42.7) | 66.2 (59.5, 73.0) |
| April | 89.2 (86.5, 91.9) | 59.2 (51.2, 67.1) | 71.1 (68.1, 74.2) |
| **<u>50-64 years</u>** |  |  |  |
| **Pfizer-BioNTech** | **95.3 (94.3, 96.2)** | **71.5 (66.5, 76.5)** | **76.2 (74.5, 78.0)** |
| January/February | 89.7 (86.3, 93.1) | 70.6 (59.7, 81.5) | 73.4 (69.3, 77.5) |
| March | 93.4 (90.7, 96.1) | 71.6 (60.9, 82.3) | 74.2 (70.1, 78.2) |
| April | 97.1 (96.2, 98.0) | 71.7 (66.0, 77.5) | 77.4 (75.4, 79.4) |
| **Moderna** | **97.4 (96.6, 98.2)** | **85.8 (82.1, 89.5)** | **82.9 (81.3, 84.6)** |
| January/February | 96.1 (94.1, 98.1) | 77.9 (69.0, 86.8) | 76.4 (72.8, 80.1) |
| March | 97.1 (95.2, 99.0) | 83.6 (75.1, 92.0) | 78.2 (74.3, 82.0) |
| April | 98.0 (97.1, 99.0) | 90.2 (86.2, 94.1) | 87.6 (85.8, 89.3) |
| **Janssen** | **86.8 (83.4, 90.2)** | **72.6 (63.4, 81.8)** | **76.0 (72.6, 79.4)** |
| March | 88.5 (82.8, 94.1) | 75.3 (59.9, 90.8) | 70.3 (63.6, 77.1) |
| April | 86.1 (81.9, 90.2) | 71.4 (60.3, 82.5) | 78.4 (74.6, 82.2) |

| ***Vaccine cohort*** | **May 1 week**<br>% VE (95% CI) | **July 10 week**<br>% VE (95% CI) | **August 28 week**<br>% VE (95% CI) |
| --- | --- | --- | --- |
| **<u>≥65 years</u>** |  |  |  |
| **Pfizer-BioNTech** | **91.9 (90.6, 93.2)** | **79.2 (74.5, 83.8)** | **77.8 (75.9, 79.6)** |
| January/February | 85.1 (81.4, 88.8) | 72.1 (61.9, 82.3) | 75.9 (72.3, 79.5) |
| March | 91.8 (89.7, 93.9) | 81.0 (74.5, 87.5) | 77.2 (74.4, 79.9) |
| April | 95.1 (93.7, 96.6) | 81.0 (75.1, 87.0) | 79.1 (76.7, 81.5) |
| **Moderna** | **96.2 (95.3, 97.0)** | **87.2 (83.8, 90.5)** | **84.3 (82.8, 85.7)** |
| January/February | 97.2 (95.4, 99.0) | 80.0 (70.1, 89.9) | 79.1 (75.2, 82.9) |
| March | 96.1 (94.8, 97.4) | 86.3 (81.4, 91.2) | 84.5 (82.5, 86.5) |
| April | 95.9 (94.7, 97.2) | 90.4 (86.4, 94.4) | 85.8 (83.9, 87.7) |
| **Janssen** | **81.7 (76.3, 87.1)** | **79.3 (68.1, 90.5)** | **70.8 (65.7, 76.0)** |
| March | 85.5 (78.3, 92.7) | 86.2 (72.6, 99.8) | 67.0 (58.9, 75.1) |
| April | 78.9 (71.3, 86.4) | 74.1 (57.7, 90.5) | 73.7 (67.4, 80.0) |

**Figure 1.**
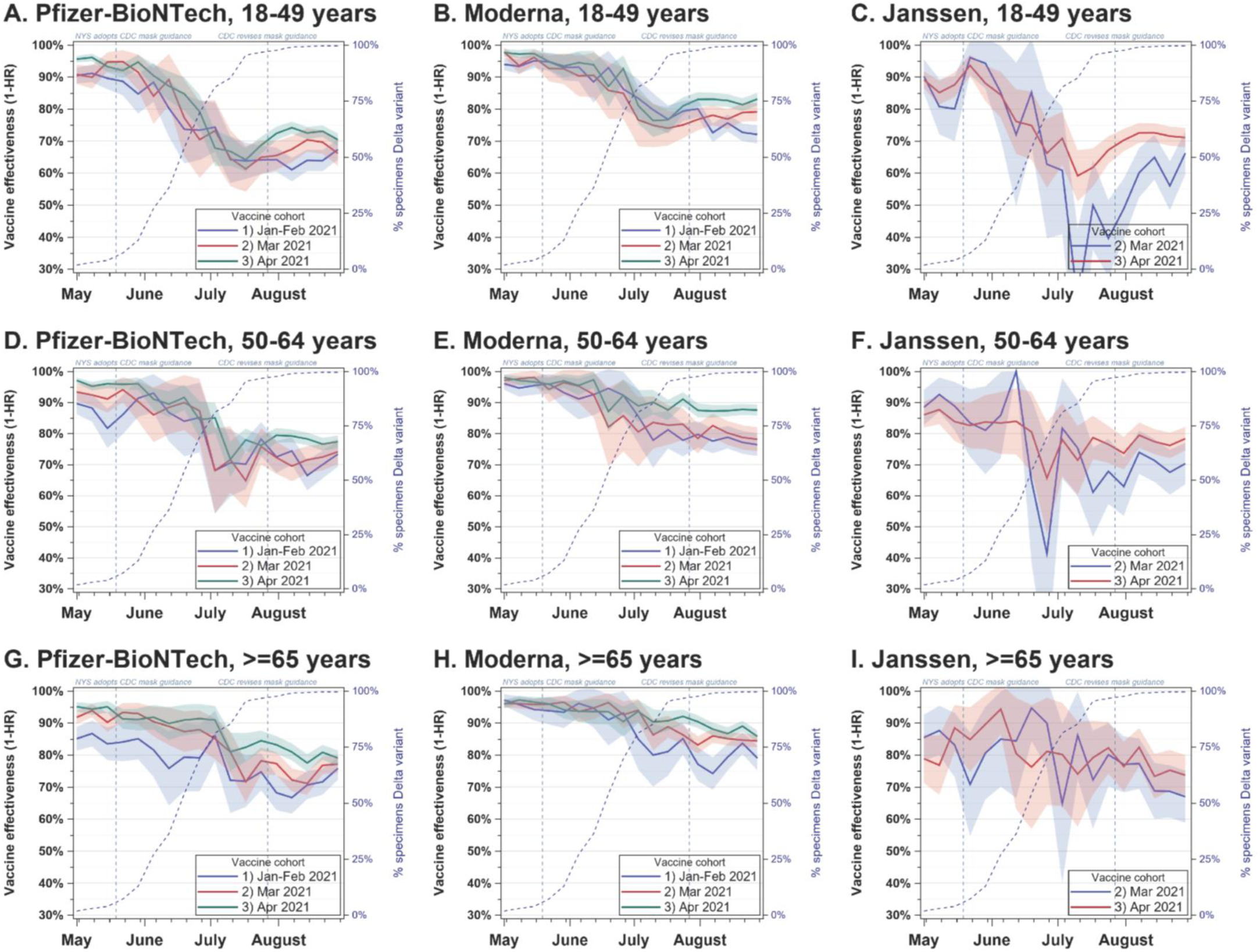
Estimated Vaccine Effectiveness for Laboratory-confirmed COVID-19 Cases, by Vaccine Product, Age, and Timing of Vaccination.

For persons 18-49 years, VE for Pfizer-BioNTech declined 27.8% from 93.6% during the May 1 to July 10 weeks, and increased 3.2% by the August 28 week (net −24.6%, *r* between logit[VE] and logit[% Delta variant] *=* −0.92, Table 1, Figure 1a-c). Similarly, Moderna VE decreased 19.3% from 96.5%, then increased 1.3% (net −18.0%, *r* = −0.94). For Janssen, VE decreased 37.7% from 89.4%, then increased 18.5% (net −19.2%, *r = -*0.76), with less stable estimates. Differences within product between time cohorts were smaller over time. At the end of observation, VE ranged 4.2% across time cohorts for Pfizer, 11.0% for Moderna, and 4.9% for Janssen.

For persons 50-64 years, VE for Pfizer-BioNTech decreased 23.8% from 95.3% during May 1 to July 10, and increased 4.7% by August 28 (net –19.1%, *r* between logit[VE] and logit[% Delta variant] *=* - 0.93, Figure 1d-f). Similarly, Moderna VE decreased 11.6% from 97.4%, then decreased 2.9% (net - 14.4%, *r* = −0.95). For Janssen, VE decreased 14.2% from 86.8%, then increased 3.4% (net −10.8%, *r* = - 0.68). At the end of observation, VE ranged 4.0% across time cohorts for Pfizer, 11.1% for Moderna, and 8.0% for Janssen, with a gradient of lower VE point estimates for earlier-vaccinated cohorts.

Among persons ≥65 years, VE for Pfizer-BioNTech decreased 12.7% from 91.9% during May 1 to July 10, and decreased 1.4% by August 28 (net −14.1%, *r* between logit[VE] and logit[% Delta variant] = - 0.95, Figure 1g-i). Likewise, Moderna VE decreased 9.0% from 96.2%, then decreased 2.9% (net −11.9%, *r* = −0.97). For Janssen, VE decreased 2.4% from 81.7%, then decreased 8.4% (net −10.9%, *r* = −0.65). At the end of observation, VE ranged 3.2% across time cohorts for Pfizer, 6.7% for Moderna, and 6.7% for Janssen, with lower VE point estimates for earlier-vaccinated cohorts.

Hospitalization rates also generally declined for all cohorts from May to June, then increased through August, with rates highest for the unvaccinated and those ≥65 years. (Figure S2). For persons in the 18-49 and 50-64 age cohorts who received Pfizer-BioNTech or Moderna, VE for hospitalization was

>90% for every month and time-cohort, except June among persons 18-49 years fully-vaccinated in April with Moderna (87.3%), with no clear time trend (Table 3, Figure 2a-f). Janssen recipients aged 18-49 had >90% VE except in June among those fully-vaccinated in March (53.7%). VE for Janssen recipients aged 50-64 ranged 86.6-93.3%.

**Table 3:** Estimated vaccine effectiveness for laboratory-confirmed COVID-19 hospitalizations.

| **Vaccine cohort** | **May 2021**<br>% VE (95% CI) | **June 2021**<br>% VE (95% CI) | **July 2021**<br>% VE (95% CI) | **August 2021**<br>% VE (95% CI) |
| --- | --- | --- | --- | --- |
| **<u>18-49 years</u>** |  |  |  |  |
| **Pfizer-BioNTech** | **96.4 (94.5, 97.7)** | **94.7 (90.8, 97.2)** | **95.9 (93.0, 97.7)** | **95.5 (94.0, 96.7)** |
| January/February | 97.9 (93.9, 99.6) | 98.3 (90.4, 100.0) | 96.3 (89.1, 99.2) | 93.1 (88.9, 96.0) |
| March | 99.0 (94.3, 100.0) | 97.4 (85.7, 99.9) | 96.3 (86.7, 99.6) | 95.2 (90.5, 97.9) |
| April | 95.1 (92.2, 97.1) | 92.5 (86.4, 96.3) | 95.6 (91.5, 98.0) | 96.6 (94.8, 97.9) |
| **Moderna** | **96.8 (94.7, 98.2)** | **92.0 (86.6, 95.5)** | **96.1 (92.9, 98.2)** | **97.5 (96.1, 98.5)** |
| January/February | 96.8 (92.6, 99.0) | 98.4 (91.1, 100.0) | 98.9 (93.6, 100.0) | 97.4 (94.6, 98.9) |
| March | 98.2 (93.5, 99.8) | 91.1 (77.1, 97.6) | 92.0 (81.2, 97.4) | 97.9 (94.6, 99.4) |
| April | 95.9 (92.0, 98.2) | 87.3 (76.5, 93.9) | 96.3 (90.6, 99.0) | 97.3 (94.9, 98.8) |
| **Janssen** | **95.9 (91.6, 98.4)** | **88.3 (76.9, 95.0)** | **94.8 (87.7, 98.3)** | **93.8 (90.2, 96.3)** |
| March | 90.7 (72.9, 98.1) | 53.7 (0.0, 83.1) | 94.4 (69.0, 99.9) | 92.7 (81.3, 98.0) |
| April | 97.1 (92.6, 99.2) | 96.4 (87.0, 99.6) | 94.8 (86.7, 98.6) | 94.1 (90.0, 96.8) |
| **<u>50-64 years</u>** |  |  |  |  |
| **Pfizer-BioNTech** | **95.8 (94.5, 96.9)** | **95.2 (92.6, 97.0)** | **93.9 (91.4, 95.8)** | **95.1 (94.1, 96.0)** |
| January/February | 93.1 (88.5, 96.1) | 93.4 (84.3, 97.9) | 96.8 (90.6, 99.3) | 94.6 (91.7, 96.7) |
| March | 94.0 (89.7, 96.8) | 96.0 (88.2, 99.2) | 91.5 (83.1, 96.4) | 95.9 (93.3, 97.7) |
| April | 97.0 (95.5, 98.0) | 95.4 (92.2, 97.5) | 93.8 (90.6, 96.0) | 95.0 (93.8, 96.1) |
| **Moderna** | **97.3 (96.1, 98.3)** | **97.0 (94.5, 98.6)** | **96.7 (94.4, 98.2)** | **96.9 (95.9, 97.7)** |
| January/February | 98.3 (95.7, 99.5) | 100.0 (95.5, 100.0) | 98.1 (93.0, 99.8) | 97.0 (94.8, 98.4) |
| March | 100.0 (98.1, 100.0) | 98.5 (91.7, 100.0) | 97.6 (91.4, 99.7) | 98.0 (95.8, 99.2) |
| April | 95.9 (93.8, 97.5) | 95.2 (90.8, 97.8) | 95.7 (92.1, 98.0) | 96.5 (95.1, 97.6) |
| **Janssen** | **88.2 (83.3, 91.8)** | **91.0 (82.6, 95.9)** | **90.4 (83.0, 95.1)** | **92.8 (90.1, 95.0)** |
| March | 90.7 (81.6, 96.0) | 93.3 (75.6, 99.2) | 86.6 (68.5, 95.7) | 92.2 (86.4, 96.0) |
| April | 87.0 (80.9, 91.6) | 90.0 (79.1, 96.0) | 92.0 (83.3, 96.8) | 93.1 (89.7, 95.5) |
| **<u>≥65 years</u>** |  |  |  |  |
| **Pfizer-BioNTech** | **95.0 (94.2, 95.7)** | **93.8 (92.3, 95.1)** | **89.6 (87.4, 91.4)** | **89.2 (88.1, 90.2)** |
| January/February | 93.2 (91.1, 94.9) | 92.8 (89.0, 95.6) | 89.3 (84.6, 92.8) | 86.2 (83.6, 88.5) |
| March | 95.8 (94.5, 96.8) | 94.1 (91.5, 96.1) | 89.0 (85.5, 91.8) | 89.6 (87.9, 91.1) |
| April | 95.2 (94.0, 96.2) | 94.1 (91.8, 95.9) | 90.1 (87.1, 92.6) | 90.3 (88.8, 91.6) |
| **Moderna** | **97.2 (96.7, 97.7)** | **96.4 (95.3, 97.4)** | **95.2 (93.9, 96.3)** | **94.1 (93.3, 94.8)** |
| January/February | 97.4 (95.7, 98.6) | 96.2 (92.5, 98.4) | 91.8 (86.7, 95.3) | 93.2 (91.0, 95.0) |
| March | 97.5 (96.6, 98.2) | 97.5 (95.9, 98.6) | 95.8 (93.8, 97.3) | 94.1 (93.0, 95.1) |
| April | 96.9 (96.0, 97.7) | 95.5 (93.4, 97.0) | 95.7 (93.7, 97.2) | 94.3 (93.2, 95.3) |
| **Janssen** | **85.5 (81.5, 88.8)** | **80.9 (72.7, 87.1)** | **82.4 (74.5, 88.4)** | **82.8 (79.0, 86.1)** |
| March | 90.4 (85.0, 94.2) | 80.5 (67.0, 89.4) | 83.0 (70.1, 91.3) | 82.9 (76.9, 87.7) |
| April | 81.8 (75.7, 86.6) | 81.2 (70.0, 88.9) | 82.0 (70.8, 89.6) | 82.7 (77.6, 86.9) |

**Figure 2.**
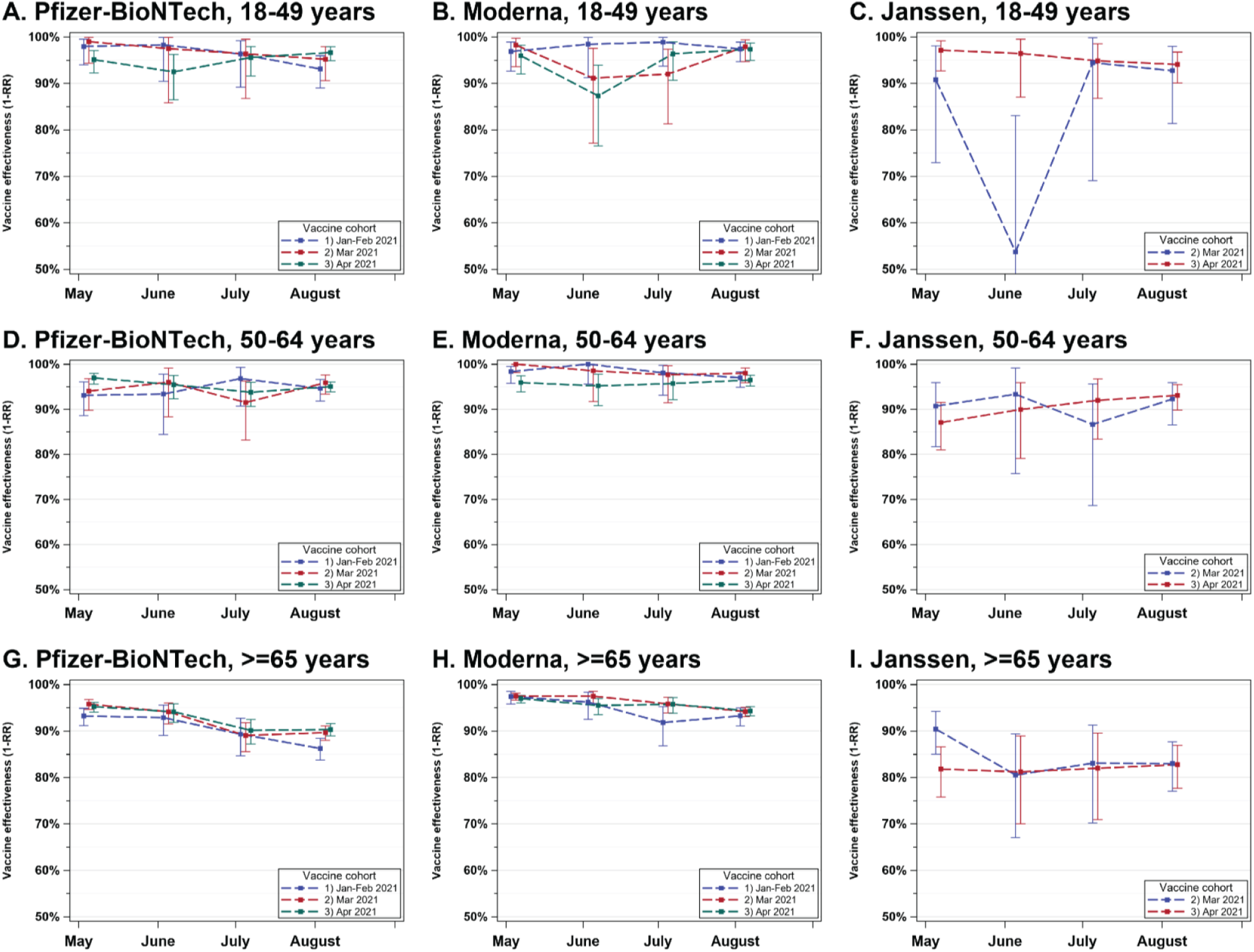
Estimated Vaccine Effectiveness for Laboratory-confirmed COVID-19 Hospitalizations, by Vaccine Product, Age, and Timing of Vaccination.

Among persons ≥65 years, hospitalization VE estimates declined for Pfizer-BioNTech recipients from May (93.2% for January/February, 95.8% for March, 95.2% for April cohorts) to August (86.2%, 89.6%, 90.3% respectively). Smaller declines were observed for Moderna recipients from May (97.4% for January/February, 96.1% for March, 95.9% for April cohorts) to August (93.2%, 94.1%, 94.3% respectively). Estimates were lower for Janssen recipients in both cohorts, ranging 80.5%-90.4%, with unclear time trend.

### Sensitivity analyses

In analyses that considered alternative population sizes for the unvaccinated, VE for cases changed medians of −3.6% when adjusting for census uncertainty and −1.6% when using the entire population size (Table S2-3). VE for hospitalization changed medians of −0.9% when adjusting for census uncertainty, −0.4% when using the entire unvaccinated population size, +2.3% when limiting to hospitalizations “for COVID-19”, and −0.4% when using the time-defined cohort sizes without exclusions applied (Table S4-7).

## Discussion

By analyzing large cohorts of NYS residents, we observed substantial declines in VE for COVID-19 cases from May to August 2021. These declines occurred simultaneously across age, product, and time-cohort, with the largest declines seen for Pfizer-BioNTech recipients. VE declines by time-cohort were not offset in calendar time by approximately one-month, which would have suggested immunologic waning. However, trends in VE were highly inversely-correlated with increasing Delta variant prevalence and plateaued for persons 18-64 years during the period in which Delta exceeded 85%. VE continued to modestly decline for persons ≥65 years. In contrast, VE for hospitalizations remained high, with lower levels for persons ≥65 years and those receiving Janssen, and modest declines over time within −6%, limited to Pfizer-BioNTech and Moderna recipients ≥65 years.

Declines in VE for COVID-19 cases during July 2021 followed by a plateau in late summer have been reported recently by multiple jurisdictions using overall rate comparisons between open cohorts.^22-24^ These observations could also reflect changes in the population vaccinated, in terms of newly-vaccinated persons entering and differences in the distribution of products received and changes in the population unvaccinated. Other recent studies have shown VE changes in pre- vs. post-Delta eras in more well-controlled designs, but with less temporal resolution and outcomes observed.^13,14^ A strength of this work is the use of closed cohorts to control for the challenges posed by previous surveillance-based approaches to demonstrate consistency in these patterns for VE. Compared to more targeted studies, this study more precisely aligns patterns in time with population-level changes in the Delta variant. The apparent lower set-point for VE after population-level saturation of the Delta variant may reflect its increased transmissibility.^25^

Policy changes co-occurred with, and were in part responding to, changes in the epidemic driven by the Delta variant and availability of vaccines. On May 19, 2021, NYS adopted the CDC guidance for fully-vaccinated persons, followed in June by the end of NYS’ state of emergency, which reduced mask-wearing, distancing and other prevention practices.^26^ On July 27, 2021, CDC recommended mask usage for fully-vaccinated persons based on transmission levels.^27^ Time-dependent changes in prevention policies and behaviors by vaccination status could contribute to observed trends; the modest VE increases observed for some groups in August may reflect such changes.

Our results provide evidence of immunity loss associated with time since completion of vaccination. For most groups, we observed gradients by time-cohort in August, whereby more recently-vaccinated cohorts had higher VE for cases, particularly for Moderna recipients, and continued declines that month for all cohorts of persons ≥65 years. Declines in VE for hospitalization of <10% were seen for persons ≥65 years, particularly those receiving Pfizer-BioNTech and in the January/February cohort. These findings are consistent with recent findings of reduced VE for severe disease in older populations, particularly those living in long-term care facilities (LTCF), and who received Pfizer.^8,12,28^ Because the federal LTCF vaccine distribution program primarily distributed Pfizer-BioNTech vaccine to NYS, the January/February cohort for Pfizer-BioNTech is more enriched for this population, potentially contributing to the decline observed in that group.

Irrespective of the cause or propensity for continued declines in VE, our findings have important implications for national vaccine policy. Pfizer-BioNTech booster doses have been demonstrated safe and to increase short-term protection against the Delta variant.^29^ Our findings align with the CDC recommendation for Pfizer-BioNTech boosters in persons ≥65 years.^16^ The booster recommendation for 18-64 years in high-exposure occupations was against the vote of ACIP members, who cited limited supportive data on VE for persons <65 years. This study addresses key U.S. data gaps, demonstrating population-wide declines in VE for infection to <85% and smaller changes in VE for severe disease limited to persons ≥65 years receiving mRNA vaccines. This suggests a focus on booster shots for those >65 years is warranted, with only limited need for booster expansion beyond that age group for the purposes of reducing severe COVID-19.

### Limitations

Our estimates of VE may be influenced by unmeasured confounding due to behavioral, medical, or exposure differences between fully-vaccinated and unvaccinated adults.^10,11^ This potential bias comes in exchange for large sample sizes and outcome numbers that exceed those of other nations’ studies and of smaller, more controlled designs.^6,7,13,14^ Our findings are further strengthened by broad age representation across time-cohorts and the consistency of changes in calendar time.

This study did not account for indirect effects (e.g. herd-immunity-type) between groups. The survival analytic approach for cases required persons to have no diagnosis 90 days before May 1, consistent with case-definitions. In this interval between Winter and Summer 2021 COVID-19 waves, under 1% of persons fully-vaccinated through April 30 received a diagnosis before or after vaccination and were excluded initially and throughout the analysis, due to the closed-cohort design. The weekly-calculated conditional hazard function was thus negligibly affected by such persons. Persons who received non-FDA authorized vaccines were excluded from the full vaccination definition and were analytically classified unvaccinated, but comprise an estimated 0.03% of persons fully vaccinated in the registries by May 2021. Finally, it is unclear the extent to which earlier SARS-CoV-2 infection modifies vaccine effectiveness; future research may explore effect modification by prior diagnostic history.^30^

### Conclusions

We observed declines in VE for cases contemporaneous with increased Delta variant prevalence, and changes in prevention policies, with little variation in timing by age, product, or time-cohort, suggesting these changes may be primarily driven by factors other than immunological waning. VE for hospitalizations remained high. Modest declines were limited to Pfizer-BioNTech and Moderna recipients ≥65 years, supporting targeted booster dosing recommendations.

## Data Availability

Data in this manuscript are not publicly available.

## Acknowledgements

Lyndsey Hoyt and Samuel Meyer, New York State Department of Health; Citywide Immunization Registry Program, New York City Department of Health and Mental Hygiene.

