## Supplementary appendix for "COVID-19 Vaccine Effectiveness by Product and Timing in New York State"

### Supplementary Methods

*Primary and sensitivity analyses*

In the primary analysis, persons with a positive laboratory result within 90 days before May 1 were considered not susceptible for either outcome and excluded, per the CDC case-definition. Sensitivity analyses probed the impact of variations in the cohort definitions.

1. The primary analysis used the 2018 Vintage Census file for defining the unvaccinated population, consistent with previous COVID-19 surveillance practice. The intercensal estimates for 2020 projected a 0.8% decrease in the NYS adult population relative to 2018, however the Decennial 2020 estimates projected a 3.9% increase, although age-specific estimates are unavailable.[31] To assess the potential impact of these added persons on the calculated unvaccinated population size, we distributed the estimated additional 606,000 adults to each age group, proportional to their 2018 representation.
2. Cases within 90 days of May 1 unmatched with the immunization registry were classified unvaccinated; a false non-match could potentially deflate the unvaccinated population. This analysis did not exclude unvaccinated persons with cases within 90 days.
3. About half of hospitalized patients in HERDS were reported admitted “for COVID-19”, using non-standardized definitions.[10] The hospitalization analysis was repeated restricted to such hospitalizations, as opposed to those “with COVID-19” in the primary analysis.
4. The primary analysis defined the susceptible population size identically for both cases and hospitalizations, using a case-based definition. Because no such susceptibility definition exists for hospitalizations, the hospitalization analyses were repeated for the entire time-defined cohort sizes without exclusions applied.

Figure S1: Weekly hazard rates for Laboratory-confirmed COVID-19 cases by Vaccine Product, Age, and Timing of Vaccination

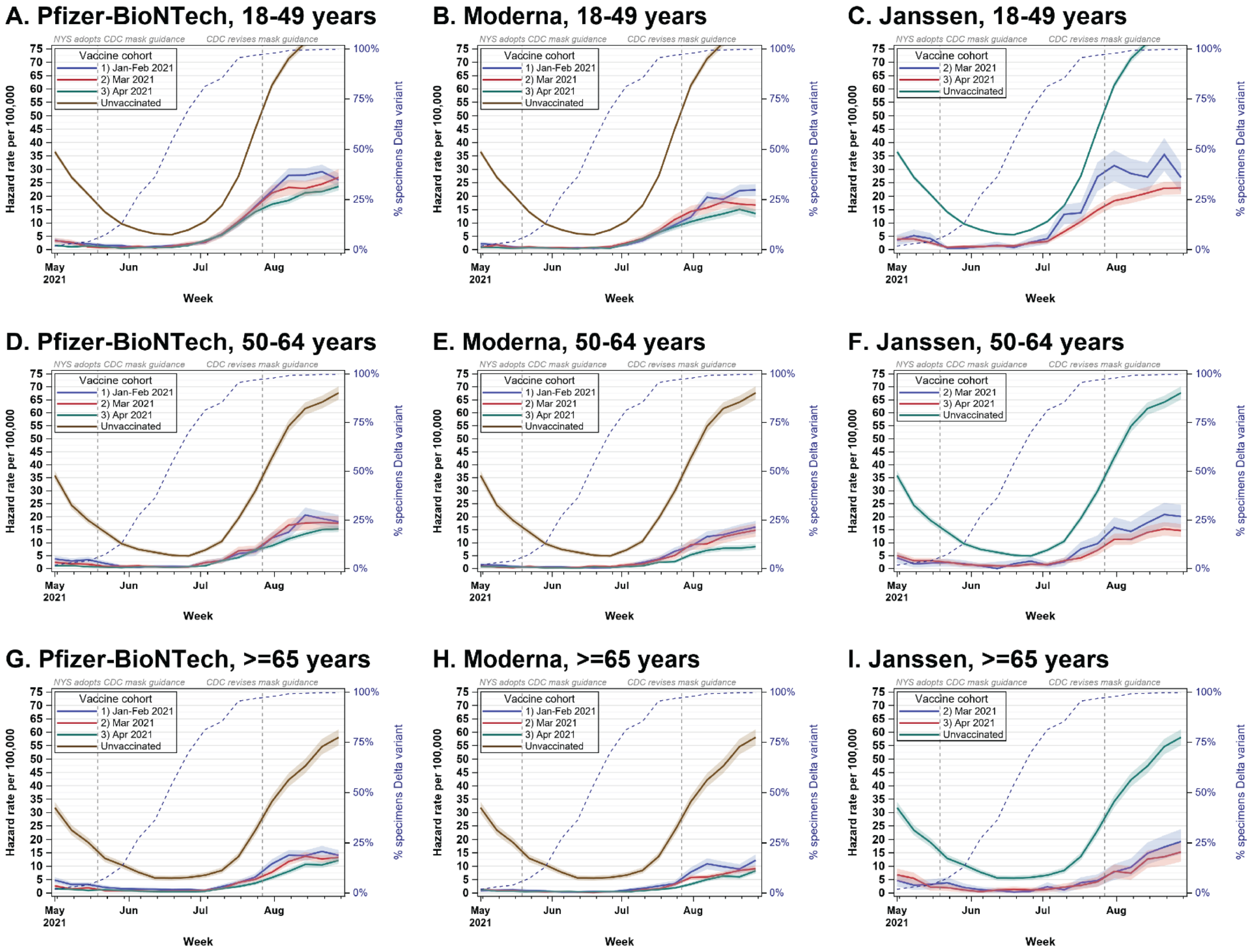

Figure S2: Monthly incidence rates for laboratory-confirmed COVID-19 hospitalization by Vaccine Product, Age, and Timing of Vaccination

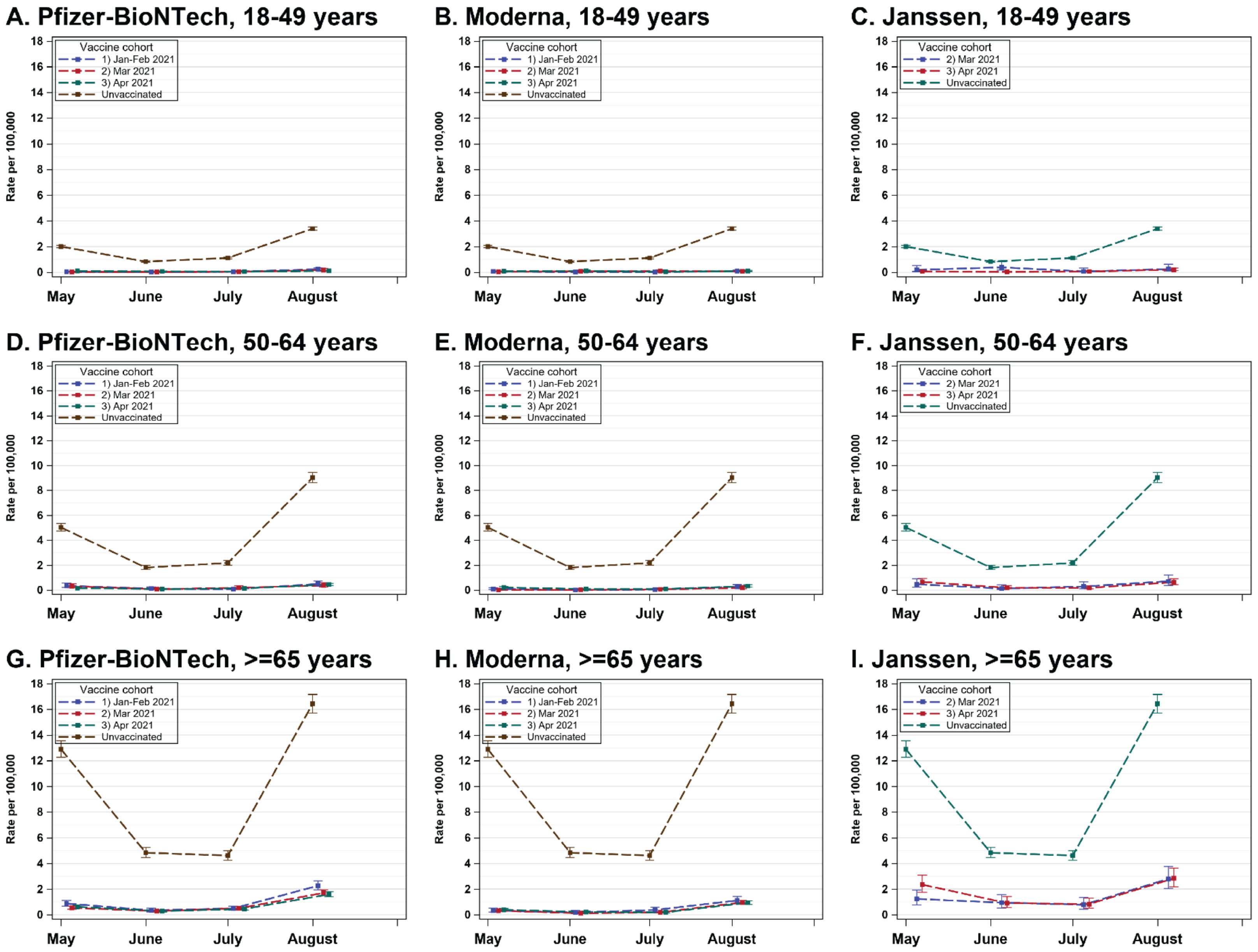

Table S1: Overview of phased vaccine eligibility in New York State

| Effective Date | Eligible Population |
|---|---|
| December 14, 2020 | High-risk hospital staff, affiliates, volunteers and contract staff, following the clinical risk assessment guidance |
| December 21, 2020 | - High-risk hospital staff including State-operated OMH psychiatric centers<br>- Emergency Medical Services (EMS) Personnel<br>- Medical Examiners and Coroners<br>- Funeral workers who have direct contact with infectious material and bodily fluids<br>- Health care or other high-risk direct care essential staff working in LTCFs and long-term, congregate settings overseen by OPWDD, OMH and OASAS<br>- Persons living in LTCFs and in long-term congregate settings overseen by OPWDD and OMH |
| December 28, 2020 | - High-risk hospital and FQHC staff, including OMH psychiatric centers<br>- Emergency Medical Services (EMS) Personnel<br>- Medical Examiners and Coroners<br>- Agency staff and residents in congregate living situations run by the Office of People with Developmental Disabilities (OPWDD) the Office of Mental Health (OMH) and the Office of Addiction Services and Supports (OASAS).<br>- Urgent Care providers<br>- Any staff administering COVID-19 Vaccinations |
| January 4, 2021 | - All Outpatient/Ambulatory front line, high risk health care providers who provide direct in-person patient care or other staff in a position where they have direct contact with patients, such as receptionists, of any age.<br>- All front line, high risk public health workers who have direct contact with patients, including those conducting COVID-19 Tests |
| January 12, 2021 | - Age 65 and older<br>- First Responder or Support Staff for First Responder Agency<br>o Fire<br>- State Fire Service, including firefighters and investigators (professional and volunteer)<br>o Local Fire Service, including firefighters and investigators (professional and volunteer)<br>- Police and Investigations<br>- State Police, including Troopers<br>- State Park Police, DEC Police, Forest Rangers<br>- SUNY Police<br>- Sheriffs' Offices<br>- County Police Departments and Police Districts<br>- City, Town, and Village Police Departments<br>- Transit of other Public Authority Police Departments |

<table>
<tr><th>Effective Date</th><th>Eligible Population</th></tr>
<tr><td>January 12, 2021<br>continued</td><td>
<ul>
<li><ul>
<li><ul><li>State Field Investigations, including DMV, SCOC, Justice Center, DFS, IG, Tax, OCFS, SLA</li></ul></li>
<li>Public Safety Communications
<ul><li>Emergency Communication and PSAP Personnel, including dispatchers and technicians</li></ul></li>
<li>Other Sworn and Civilian Personnel
<ul><li>Court Officer</li><li>Other Police or Peace Officer</li><li>Support or Civilian Staff for Any of the Above Services, Agencies, or Facilities</li></ul></li>
</ul></li>
<li>Corrections
<ul><li>State DOCCS Personnel, including correction and parole officers</li><li>Local Correctional Facilities, including correction officers</li><li>Local Probation Departments, including probation officers</li><li>State Juvenile Detention and Rehabilitation Facilities</li><li>Local Juvenile Detention and Rehabilitation Facilities</li></ul></li>
<li>P-12 Schools
<ul><li>P-12 school (public or non-public) or school district faculty or staff (includes all teachers, substitute teachers, student teachers, school administrators, paraprofessional staff, and support staff including bus drivers)</li><li>Contractor working in a P-12 school (public or non-public) or school district (including contracted bus drivers)</li><li>Licensed, registered, approved or legally exempt group childcare</li></ul></li>
<li>In-Person College Faculty and Instructors</li>
<li>Employees or Support Staff of Licensed, Registered, Approved or Legally Exempt Group Childcare Settings</li>
<li>Licensed, Registered, approved or legally exempt group Childcare Provider</li>
<li>Public Transit
<ul><li>Airline and airport employee</li><li>Passenger railroad employee</li><li>Subway and mass transit employee (i.e., MTA, LIRR, Metro North, NYC Transit, Upstate transit)</li><li>Ferry employee</li><li>Port Authority employee</li><li>Public bus employee</li></ul></li>
<li>Public Facing Grocery Store Workers</li>
<li>Individual living in a homeless shelter where sleeping, bathing or eating accommodations must be shared with individuals and families who are not part of your household.</li>
<li>Individual working (paid or unpaid) in a homeless shelter where sleeping, bathing or eating accommodations must be shared by individuals and families who are not part of the same household, in a position where there is potential for interaction with shelter residents.</li>
</ul>
</td></tr>
</table>

| Effective Date | Eligible Population |
|---|---|
| February 8, 2021 | - Restaurant employees,<br>- Restaurant delivery workers, and<br>- For-hire vehicle drivers, including taxi, livery, black car, and transportation network company drivers |
| February 15, 2021 | **Individuals with comorbidities and underlying conditions are eligible to receive COVID-19 vaccine.** The list is subject to change as additional scientific evidence is published and as New York State obtains and analyzes additional state-specific data. Adults over the age of 16 with the following conditions due to increased risk of moderate or severe illness or death from the virus that causes COVID-19 are eligible:<br>• Cancer (current or in remission, including 9/11-related cancers);<br>• Chronic kidney disease;<br>• Pulmonary Disease, including but not limited to, COPD (chronic obstructive pulmonary disease), asthma (moderate-to-severe), pulmonary fibrosis, cystic fibrosis, and 9/11 related pulmonary diseases;<br>• Intellectual and Developmental Disabilities including Down Syndrome;<br>• Heart conditions, including but not limited to heart failure, coronary artery disease, cardiomyopathies, or hypertension (high blood pressure);<br>• Immunocompromised state (weakened immune system) including but not limited to solid organ transplant or from blood or bone marrow transplant, immune deficiencies, HIV, use of corticosteroids, use of other immune weakening medicines, or other causes;<br>• Severe Obesity (BMI 40 kg/m2), Obesity (body mass index [BMI] of 30 kg/m2 or higher but < 40 kg/m2);<br>• Pregnancy;<br>• Sickle cell disease or Thalassemia;<br>• Type 1 or 2 diabetes mellitus;<br>• Cerebrovascular disease (affects blood vessels and blood supply to the brain);<br>• Neurologic conditions, including but not limited to Alzheimer's Disease or dementia; and<br>• Liver disease. |
| March 1, 2021 | - Public-facing hotel workers. |
| March 17, 2021 | - Public-facing government and public employees,<br>- Not-for-profit workers who provide public-facing services to New Yorkers in need, and<br>- Essential in-person public-facing building service workers and providers of essential building services. |
| March 23, 2021 | - New York Residents age 50 and older. |
| March 30, 2021 | - New York Residents age 30 to 50. |
| April 6, 2021 | - All New York State residents age 16 and older, including individuals studying in New York, or individuals employed in the State of New York |
| May 6, 2021 | - All individuals 16 years of age and older that reside in the United States |
| May 13, 2021 | - All individuals **12 years of age and older** that reside in the United States |

Table S2: Estimated vaccine effectiveness for laboratory-confirmed COVID-19 cases, adjusted for census uncertainty (sensitivity analysis #1)

| ***Vaccine cohort*** | **May 1 week**<br>*% VE (95% CI)* | **July 10 week**<br>% VE (95% CI) | **August 28 week**<br>% VE (95% CI) |
|---|---|---|---|
| **18-49 years** | | | |
| **Pfizer-BioNTech** | **92.5 (91.3, 93.7)** | **59.9 (55.6, 64.3)** | **63.5 (61.6, 65.4)** |
| January/February | 88.8 (85.9, 91.8) | 58.0 (49.5, 66.6) | 61.6 (57.8, 65.3) |
| March | 89.3 (85.8, 92.7) | 58.5 (48.2, 68.8) | 60.2 (55.5, 64.8) |
| April | 94.9 (93.6, 96.1) | 61.1 (55.7, 66.4) | 65.2 (62.9, 67.4) |
| **Moderna** | **95.9 (94.9, 96.8)** | **73.2 (69.3, 77.1)** | **74.6 (72.9, 76.3)** |
| January/February | 93.0 (90.8, 95.2) | 76.3 (70.1, 82.4) | 67.1 (63.8, 70.4) |
| March | 97.5 (95.9, 99.0) | 70.6 (62.5, 78.7) | 75.4 (72.1, 78.8) |
| April | 97.3 (96.0, 98.5) | 72.3 (66.3, 78.3) | 80.1 (77.8, 82.4) |
| **Janssen** | **87.6 (84.7, 90.4)** | **43.4 (34.1, 52.6)** | **64.9 (61.6, 68.2)** |
| March | 88.6 (82.4, 94.8) | 5.9 (0.0, 32.8) | 60.2 (52.3, 68.2) |
| April | 87.3 (84.2, 90.5) | 52.1 (42.7, 61.5) | 66.0 (62.4, 69.6) |
| **50-64 years** | | | |
| **Pfizer-BioNTech** | **94.5 (93.4, 95.6)** | **66.6 (60.8, 72.4)** | **72.0 (69.9, 74.1)** |
| January/February | 87.9 (83.9, 91.9) | 65.5 (52.8, 78.3) | 68.7 (63.9, 73.5) |
| March | 92.3 (89.1, 95.5) | 66.7 (54.2, 79.2) | 69.6 (64.9, 74.3) |
| April | 96.6 (95.6, 97.7) | 66.8 (60.1, 73.6) | 73.4 (71.1, 75.7) |
| **Moderna** | **96.9 (96.0, 97.9)** | **83.4 (79.0, 87.7)** | **79.9 (78.0, 81.8)** |
| January/February | 95.4 (93.1, 97.8) | 74.1 (63.6, 84.5) | 72.3 (68.0, 76.5) |
| March | 96.6 (94.4, 98.8) | 80.7 (70.8, 90.7) | 74.3 (69.8, 78.9) |
| April | 97.7 (96.6, 98.8) | 88.4 (83.8, 93.1) | 85.4 (83.3, 87.5) |
| **Janssen** | **84.5 (80.6, 88.5)** | **67.9 (57.1, 78.6)** | **71.7 (67.7, 75.7)** |
| March | 86.5 (79.8, 93.1) | 71.1 (52.9, 89.2) | 65.1 (57.2, 73.0) |
| April | 83.7 (78.8, 88.5) | 66.5 (53.4, 79.5) | 74.6 (70.1, 79.0) |
| **≥65 years** | | | |
| **Pfizer-BioNTech** | **90.5 (88.9, 92.0)** | **75.6 (70.1, 81.0)** | **73.8 (71.7, 76.0)** |
| January/February | 82.6 (78.2, 86.9) | 67.3 (55.4, 79.3) | 71.7 (67.4, 75.9) |
| March | 90.4 (87.9, 92.8) | 77.7 (70.1, 85.3) | 73.2 (69.9, 76.4) |
| April | 94.3 (92.6, 96.0) | 77.8 (70.8, 84.7) | 75.4 (72.6, 78.2) |
| **Moderna** | **95.5 (94.5, 96.5)** | **84.9 (81.0, 88.9)** | **81.5 (79.8, 83.2)** |
| January/February | 96.7 (94.6, 98.9) | 76.5 (65.0, 88.1) | 75.4 (70.8, 79.9) |
| March | 95.4 (93.9, 96.9) | 83.9 (78.2, 89.7) | 81.8 (79.4, 84.1) |
| April | 95.2 (93.7, 96.8) | 88.7 (84.0, 93.4) | 83.3 (81.1, 85.5) |
| **Janssen** | **78.6 (72.3, 84.9)** | **75.7 (62.6, 88.8)** | **65.7 (59.7, 71.7)** |
| March | 83.0 (74.6, 91.4) | 83.8 (67.8, 99.8) | 61.2 (51.6, 70.8) |
| April | 75.3 (66.4, 84.1) | 69.6 (50.3, 88.9) | 69.1 (61.7, 76.5) |

Table S3: Estimated vaccine effectiveness for laboratory-confirmed COVID-19 cases, using the entire unvaccinated population size (sensitivity analysis #2)

| ***Vaccine cohort*** | **May 1 week**<br>*% VE (95% CI)* | **July 10 week**<br>% VE (95% CI) | **August 28 week**<br>% VE (95% CI) |
|---|---|---|---|
| **18-49 years** | | | |
| **Pfizer-BioNTech** | **93.1 (92.0, 94.2)** | **63.2 (59.2, 67.2)** | **66.5 (64.8, 68.3)** |
| January/February | 89.7 (87.1, 92.4) | 61.4 (53.6, 69.3) | 64.8 (61.4, 68.2) |
| March | 90.1 (86.9, 93.3) | 61.9 (52.4, 71.4) | 63.5 (59.2, 67.7) |
| April | 95.3 (94.1, 96.4) | 64.2 (59.3, 69.1) | 68.1 (65.9, 70.2) |
| **Moderna** | **96.2 (95.3, 97.1)** | **75.4 (71.8, 79.0)** | **76.7 (75.1, 78.3)** |
| January/February | 93.5 (91.5, 95.6) | 78.2 (72.6, 83.8) | 69.9 (66.8, 72.9) |
| March | 97.7 (96.2, 99.1) | 73.0 (65.6, 80.4) | 77.5 (74.4, 80.6) |
| April | 97.5 (96.3, 98.6) | 74.5 (69.1, 80.0) | 81.8 (79.7, 83.9) |
| **Janssen** | **88.6 (86.0, 91.2)** | **48.0 (39.5, 56.5)** | **67.8 (64.8, 70.8)** |
| March | 89.5 (83.8, 95.2) | 13.5 (0.0, 38.3) | 63.5 (56.2, 70.8) |
| April | 88.3 (85.4, 91.3) | 56.0 (47.4, 64.6) | 68.8 (65.5, 72.1) |
| **50-64 years** | | | |
| **Pfizer-BioNTech** | **94.9 (93.9, 95.9)** | **69.1 (63.7, 74.5)** | **74.2 (72.3, 76.1)** |
| January/February | 88.8 (85.1, 92.5) | 68.1 (56.3, 79.9) | 71.1 (66.7, 75.5) |
| March | 92.9 (89.9, 95.8) | 69.2 (57.6, 80.8) | 71.9 (67.6, 76.3) |
| April | 96.9 (95.9, 97.8) | 69.3 (63.1, 75.6) | 75.5 (73.3, 77.6) |
| **Moderna** | **97.2 (96.3, 98.0)** | **84.6 (80.6, 88.6)** | **81.5 (79.7, 83.2)** |
| January/February | 95.8 (93.6, 97.9) | 76.0 (66.3, 85.7) | 74.4 (70.4, 78.3) |
| March | 96.9 (94.8, 98.9) | 82.2 (73.0, 91.4) | 76.3 (72.1, 80.5) |
| April | 97.9 (96.9, 98.9) | 89.3 (85.0, 93.6) | 86.5 (84.6, 88.4) |
| **Janssen** | **85.7 (82.0, 89.3)** | **70.3 (60.3, 80.3)** | **73.9 (70.2, 77.6)** |
| March | 87.5 (81.3, 93.7) | 73.3 (56.5, 90.0) | 67.8 (60.5, 75.1) |
| April | 84.9 (80.4, 89.4) | 69.0 (57.0, 81.1) | 76.5 (72.4, 80.6) |
| **≥65 years** | | | |
| **Pfizer-BioNTech** | **91.2 (89.8, 92.6)** | **77.5 (72.5, 82.5)** | **75.9 (73.9, 77.9)** |
| January/February | 83.9 (79.9, 87.9) | 69.9 (58.8, 80.9) | 73.9 (70.0, 77.8) |
| March | 91.1 (88.9, 93.4) | 79.4 (72.4, 86.4) | 75.3 (72.3, 78.3) |
| April | 94.7 (93.2, 96.3) | 79.5 (73.1, 85.9) | 77.3 (74.7, 79.9) |
| **Moderna** | **95.9 (95.0, 96.8)** | **86.1 (82.5, 89.7)** | **83.0 (81.4, 84.6)** |
| January/February | 97.0 (95.0, 99.0) | 78.4 (67.7, 89.0) | 77.3 (73.1, 81.5) |
| March | 95.8 (94.4, 97.1) | 85.2 (79.9, 90.5) | 83.2 (81.0, 85.4) |
| April | 95.6 (94.2, 97.0) | 89.6 (85.3, 93.9) | 84.6 (82.6, 86.7) |
| **Janssen** | **80.2 (74.4, 86.0)** | **77.6 (65.5, 89.7)** | **68.4 (62.9, 74.0)** |
| March | 84.3 (76.5, 92.1) | 85.1 (70.3, 99.8) | 64.3 (55.4, 73.1) |
| April | 77.2 (69.0, 85.3) | 72.0 (54.2, 89.7) | 71.5 (64.7, 78.4) |

Table S4: Estimated vaccine effectiveness for laboratory-confirmed COVID-19 hospitalizations, adjusted for census uncertainty (sensitivity analysis #1)

| *Vaccine cohort* | **May 2021** *% VE (95% CI)* | **June 2021** % VE (95% CI) | **July 2021** % VE (95% CI) | **August 2021** % VE (95% CI) |
|---|---|---|---|---|
| **18-49 years** | | | | |
| **Pfizer-BioNTech** | **95.8 (93.6, 97.4)** | **93.7 (89.2, 96.7)** | **95.2 (91.8, 97.4)** | **94.8 (93.0, 96.2)** |
| January/February | 97.6 (92.9, 99.5) | 98.0 (88.7, 99.9) | 95.6 (87.2, 99.1) | 91.9 (87.0, 95.3) |
| March | 98.8 (93.3, 100.0) | 97.0 (83.3, 99.9) | 95.7 (84.4, 99.5) | 94.4 (88.9, 97.6) |
| April | 94.2 (90.8, 96.6) | 91.2 (84.1, 95.6) | 94.8 (90.1, 97.6) | 96.0 (93.9, 97.5) |
| **Moderna** | **96.2 (93.8, 97.9)** | **90.6 (84.3, 94.8)** | **95.5 (91.7, 97.8)** | **97.0 (95.4, 98.2)** |
| January/February | 96.3 (91.3, 98.8) | 98.1 (89.6, 100.0) | 98.7 (92.5, 100.0) | 96.9 (93.7, 98.8) |
| March | 97.9 (92.4, 99.7) | 89.6 (73.1, 97.2) | 90.6 (78.0, 97.0) | 97.5 (93.7, 99.3) |
| April | 95.2 (90.6, 97.9) | 85.1 (72.4, 92.9) | 95.7 (89.0, 98.8) | 96.8 (94.0, 98.6) |
| **Janssen** | **95.2 (90.1, 98.1)** | **86.4 (72.9, 94.1)** | **93.9 (85.6, 98.0)** | **92.8 (88.5, 95.7)** |
| March | 89.2 (68.2, 97.8) | 45.8 (0.0, 80.2) | 93.5 (63.7, 99.8) | 91.5 (78.1, 97.7) |
| April | 96.6 (91.4, 99.1) | 95.8 (84.7, 99.5) | 93.9 (84.4, 98.4) | 93.0 (88.3, 96.2) |
| **50-64 years** | | | | |
| **Pfizer-BioNTech** | **95.1 (93.6, 96.4)** | **94.4 (91.3, 96.5)** | **92.8 (89.9, 95.1)** | **94.3 (93.1, 95.3)** |
| January/February | 91.9 (86.6, 95.5) | 92.2 (81.7, 97.5) | 96.3 (89.0, 99.2) | 93.7 (90.3, 96.1) |
| March | 93.0 (87.9, 96.3) | 95.3 (86.2, 99.0) | 90.0 (80.2, 95.7) | 95.2 (92.2, 97.3) |
| April | 96.4 (94.8, 97.7) | 94.7 (90.9, 97.1) | 92.7 (89.0, 95.4) | 94.2 (92.7, 95.4) |
| **Moderna** | **96.9 (95.4, 98.0)** | **96.5 (93.5, 98.4)** | **96.1 (93.4, 97.9)** | **96.4 (95.3, 97.3)** |
| January/February | 98.0 (95.0, 99.5) | 100.0 (94.8, 100.0) | 97.8 (91.8, 99.7) | 96.5 (93.9, 98.1) |
| March | 100.0 (97.8, 100.0) | 98.3 (90.2, 100.0) | 97.2 (89.9, 99.7) | 97.6 (95.1, 99.1) |
| April | 95.2 (92.7, 97.0) | 94.4 (89.2, 97.4) | 95.0 (90.7, 97.6) | 95.9 (94.2, 97.2) |
| **Janssen** | **86.1 (80.5, 90.5)** | **89.4 (79.7, 95.2)** | **88.7 (80.1, 94.2)** | **91.6 (88.4, 94.1)** |
| March | 89.1 (78.5, 95.3) | 92.2 (71.5, 99.1) | 84.3 (63.1, 94.9) | 90.9 (84.1, 95.3) |
| April | 84.8 (77.6, 90.1) | 88.2 (75.5, 95.3) | 90.6 (80.4, 96.2) | 91.9 (88.0, 94.8) |
| **≥65 years** | | | | |
| **Pfizer-BioNTech** | **94.1 (93.2, 95.0)** | **92.8 (90.9, 94.3)** | **87.8 (85.3, 89.9)** | **87.4 (86.1, 88.6)** |
| January/February | 92.0 (89.6, 94.0) | 91.6 (87.1, 94.9) | 87.4 (82.0, 91.6) | 83.9 (80.9, 86.5) |
| March | 95.1 (93.6, 96.3) | 93.1 (90.0, 95.4) | 87.1 (83.0, 90.4) | 87.8 (85.9, 89.6) |
| April | 94.4 (93.0, 95.5) | 93.1 (90.4, 95.2) | 88.4 (84.9, 91.3) | 88.7 (86.9, 90.2) |
| **Moderna** | **96.8 (96.1, 97.4)** | **95.8 (94.5, 96.9)** | **94.4 (92.8, 95.7)** | **93.1 (92.2, 93.9)** |
| January/February | 97.0 (95.0, 98.3) | 95.5 (91.2, 98.1) | 90.4 (84.5, 94.4) | 92.1 (89.5, 94.1) |
| March | 97.0 (96.0, 97.9) | 97.0 (95.1, 98.3) | 95.1 (92.8, 96.8) | 93.1 (91.8, 94.3) |
| April | 96.4 (95.3, 97.3) | 94.7 (92.3, 96.5) | 95.0 (92.6, 96.7) | 93.3 (92.0, 94.4) |
| **Janssen** | **83.0 (78.3, 86.9)** | **77.6 (68.1, 84.9)** | **79.4 (70.1, 86.4)** | **79.9 (75.4, 83.7)** |
| March | 88.8 (82.4, 93.2) | 77.2 (61.4, 87.6) | 80.1 (65.0, 89.8) | 80.0 (73.0, 85.6) |
| April | 78.7 (71.6, 84.3) | 78.0 (64.8, 87.0) | 78.9 (65.8, 87.8) | 79.8 (73.7, 84.7) |

Table S5: Estimated vaccine effectiveness for laboratory-confirmed COVID-19 hospitalizations, using the entire unvaccinated population size (sensitivity analysis #2)

| *Vaccine cohort* | **May 2021** *% VE (95% CI)* | **June 2021** % VE (95% CI) | **July 2021** % VE (95% CI) | **August 2021** % VE (95% CI) |
|---|---|---|---|---|
| **18-49 years** | | | | |
| **Pfizer-BioNTech** | **96.1 (94.1, 97.6)** | **94.2 (90.1, 97.0)** | **95.5 (92.5, 97.6)** | **95.2 (93.6, 96.5)** |
| January/February | 97.8 (93.5, 99.5) | 98.1 (89.6, 100.0) | 96.0 (88.3, 99.2) | 92.6 (88.1, 95.7) |
| March | 98.9 (93.9, 100.0) | 97.3 (84.6, 99.9) | 96.0 (85.7, 99.5) | 94.8 (89.8, 97.8) |
| April | 94.7 (91.6, 96.9) | 91.9 (85.4, 96.0) | 95.2 (90.9, 97.8) | 96.3 (94.4, 97.7) |
| **Moderna** | **96.5 (94.3, 98.1)** | **91.4 (85.6, 95.2)** | **95.8 (92.3, 98.0)** | **97.3 (95.8, 98.3)** |
| January/February | 96.6 (92.0, 98.9) | 98.3 (90.5, 100.0) | 98.8 (93.1, 100.0) | 97.2 (94.2, 98.9) |
| March | 98.1 (93.1, 99.8) | 90.4 (75.3, 97.4) | 91.4 (79.8, 97.2) | 97.7 (94.2, 99.4) |
| April | 95.6 (91.3, 98.1) | 86.3 (74.6, 93.5) | 96.1 (89.9, 98.9) | 97.1 (94.5, 98.7) |
| **Janssen** | **95.6 (90.9, 98.2)** | **87.5 (75.1, 94.6)** | **94.4 (86.8, 98.2)** | **93.3 (89.4, 96.1)** |
| March | 90.0 (70.8, 97.9) | 50.2 (-9.2, 81.8) | 94.0 (66.6, 99.8) | 92.2 (79.9, 97.9) |
| April | 96.9 (92.1, 99.2) | 96.1 (86.0, 99.5) | 94.4 (85.7, 98.5) | 93.6 (89.2, 96.5) |
| **50-64 years** | | | | |
| **Pfizer-BioNTech** | **95.5 (94.1, 96.6)** | **94.8 (92.0, 96.8)** | **93.4 (90.6, 95.5)** | **94.7 (93.6, 95.6)** |
| January/February | 92.5 (87.6, 95.8) | 92.8 (83.0, 97.7) | 96.5 (89.8, 99.3) | 94.2 (91.0, 96.4) |
| March | 93.5 (88.8, 96.6) | 95.7 (87.3, 99.1) | 90.8 (81.7, 96.0) | 95.5 (92.8, 97.5) |
| April | 96.7 (95.2, 97.8) | 95.1 (91.6, 97.3) | 93.2 (89.8, 95.7) | 94.6 (93.3, 95.8) |
| **Moderna** | **97.1 (95.7, 98.1)** | **96.8 (94.0, 98.5)** | **96.4 (93.9, 98.1)** | **96.7 (95.6, 97.5)** |
| January/February | 98.2 (95.4, 99.5) | 100.0 (95.2, 100.0) | 97.9 (92.4, 99.7) | 96.7 (94.4, 98.3) |
| March | 100.0 (97.9, 100.0) | 98.4 (91.0, 100.0) | 97.4 (90.6, 99.7) | 97.8 (95.5, 99.1) |
| April | 95.6 (93.3, 97.2) | 94.8 (90.0, 97.6) | 95.4 (91.4, 97.8) | 96.2 (94.7, 97.4) |
| **Janssen** | **87.2 (81.9, 91.2)** | **90.2 (81.2, 95.6)** | **89.6 (81.6, 94.7)** | **92.2 (89.2, 94.5)** |
| March | 89.9 (80.1, 95.7) | 92.8 (73.6, 99.1) | 85.5 (65.9, 95.3) | 91.6 (85.3, 95.7) |
| April | 86.0 (79.3, 90.9) | 89.1 (77.3, 95.7) | 91.3 (81.9, 96.5) | 92.5 (88.9, 95.2) |
| **≥65 years** | | | | |
| **Pfizer-BioNTech** | **94.6 (93.7, 95.4)** | **93.3 (91.6, 94.7)** | **88.7 (86.4, 90.7)** | **88.4 (87.2, 89.4)** |
| January/February | 92.6 (90.4, 94.5) | 92.3 (88.1, 95.3) | 88.4 (83.3, 92.2) | 85.1 (82.3, 87.5) |
| March | 95.4 (94.1, 96.5) | 93.6 (90.8, 95.7) | 88.1 (84.3, 91.2) | 88.8 (86.9, 90.4) |
| April | 94.8 (93.5, 95.9) | 93.6 (91.1, 95.6) | 89.3 (86.1, 91.9) | 89.5 (87.9, 91.0) |
| **Moderna** | **97.0 (96.4, 97.6)** | **96.1 (94.9, 97.1)** | **94.8 (93.4, 96.0)** | **93.6 (92.8, 94.3)** |
| January/February | 97.2 (95.4, 98.4) | 95.9 (91.8, 98.2) | 91.1 (85.7, 94.9) | 92.7 (90.3, 94.6) |
| March | 97.3 (96.3, 98.0) | 97.3 (95.5, 98.4) | 95.5 (93.3, 97.1) | 93.7 (92.4, 94.7) |
| April | 96.7 (95.7, 97.5) | 95.1 (92.9, 96.8) | 95.4 (93.2, 97.0) | 93.8 (92.6, 94.9) |
| **Janssen** | **84.3 (80.0, 87.9)** | **79.4 (70.5, 86.0)** | **81.0 (72.4, 87.4)** | **81.4 (77.3, 85.0)** |
| March | 89.6 (83.7, 93.8) | 78.9 (64.3, 88.6) | 81.7 (67.7, 90.6) | 81.6 (75.1, 86.7) |
| April | 80.3 (73.7, 85.5) | 79.7 (67.5, 88.0) | 80.5 (68.5, 88.7) | 81.3 (75.8, 85.9) |

### Table S6: Estimated vaccine effectiveness for laboratory-confirmed COVID-19 hospitalizations, limiting to hospitalizations reported 'for COVID-19' (sensitivity analysis #3) [1]

| ***Vaccine cohort*** | **May 2021**<br>*% VE (95% CI)* | **June 2021**<br>% VE (95% CI) | **July 2021**<br>% VE (95% CI) | **August 2021**<br>% VE (95% CI) |
|---|---|---|---|---|
| **18-49 years** | | | | |
| **Pfizer-BioNTech** | **99.0 (97.8, 99.6)** | **97.5 (94.6, 99.1)** | **98.2 (96.1, 99.4)** | **98.1 (97.0, 98.8)** |
| January/February | 100.0 (97.5, 100.0) | 100.0 (93.6, 100.0) | 97.5 (91.0, 99.7) | 96.7 (93.6, 98.6) |
| March | 100.0 (96.2, 100.0) | 97.4 (85.7, 99.9) | 96.3 (86.7, 99.6) | 99.4 (96.6, 100.0) |
| April | 98.4 (96.4, 99.4) | 96.6 (92.0, 98.9) | 99.0 (96.4, 99.9) | 98.2 (96.8, 99.1) |
| **Moderna** | **98.9 (97.5, 99.7)** | **97.9 (94.5, 99.4)** | **97.7 (94.9, 99.2)** | **99.0 (98.0, 99.6)** |
| January/February | 99.4 (96.5, 100.0) | 100.0 (94.1, 100.0) | 98.9 (93.6, 100.0) | 99.3 (97.3, 99.9) |
| March | 99.1 (95.0, 100.0) | 95.5 (83.8, 99.5) | 93.6 (83.5, 98.3) | 98.9 (96.2, 99.9) |
| April | 98.5 (95.5, 99.7) | 97.5 (90.8, 99.7) | 99.1 (94.9, 100.0) | 98.8 (96.9, 99.7) |
| **Janssen** | **98.3 (94.9, 99.6)** | **95.6 (87.2, 99.1)** | **95.8 (89.2, 98.9)** | **96.2 (93.2, 98.1)** |
| March | 96.9 (82.8, 99.9) | 84.6 (44.0, 98.1) | 94.4 (69.0, 99.9) | 98.2 (89.8, 100.0) |
| April | 98.6 (94.8, 99.8) | 98.2 (89.9, 100.0) | 96.1 (88.6, 99.2) | 95.8 (92.2, 98.0) |
| **50-64 years** | | | | |
| **Pfizer-BioNTech** | **98.2 (97.3, 98.8)** | **99.3 (98.1, 99.9)** | **96.2 (94.1, 97.6)** | **97.0 (96.2, 97.7)** |
| January/February | 94.9 (90.9, 97.5) | 98.7 (92.5, 100.0) | 98.9 (94.0, 100.0) | 95.6 (93.0, 97.5) |
| March | 97.2 (94.0, 99.0) | 100.0 (95.1, 100.0) | 93.6 (86.0, 97.7) | 97.9 (95.9, 99.1) |
| April | 99.2 (98.4, 99.7) | 99.3 (97.6, 99.9) | 96.1 (93.5, 97.8) | 97.2 (96.2, 97.9) |
| **Moderna** | **98.9 (98.0, 99.4)** | **98.8 (96.9, 99.7)** | **98.8 (97.2, 99.6)** | **98.6 (97.9, 99.1)** |
| January/February | 100.0 (98.5, 100.0) | 100.0 (95.5, 100.0) | 100.0 (96.4, 100.0) | 98.6 (97.0, 99.5) |
| March | 100.0 (98.1, 100.0) | 100.0 (94.5, 100.0) | 98.8 (93.3, 100.0) | 99.1 (97.5, 99.8) |
| April | 98.0 (96.3, 99.0) | 97.9 (94.5, 99.4) | 98.3 (95.6, 99.5) | 98.4 (97.4, 99.1) |
| **Janssen** | **92.3 (88.3, 95.2)** | **96.0 (89.6, 98.9)** | **91.2 (84.0, 95.6)** | **95.0 (92.6, 96.7)** |
| March | 94.2 (86.4, 98.1) | 96.7 (81.2, 99.9) | 86.6 (68.5, 95.7) | 93.5 (88.1, 96.9) |
| April | 91.5 (86.4, 95.1) | 95.7 (87.3, 99.1) | 93.1 (84.9, 97.5) | 95.6 (92.8, 97.5) |
| **≥65 years** | | | | |
| **Pfizer-BioNTech** | **97.7 (97.1, 98.1)** | **97.9 (97.0, 98.6)** | **94.7 (93.2, 95.9)** | **93.4 (92.5, 94.1)** |
| January/February | 97.0 (95.6, 98.1) | 97.6 (95.1, 99.0) | 95.8 (92.7, 97.9) | 91.3 (89.2, 93.0) |
| March | 98.0 (97.1, 98.7) | 97.2 (95.3, 98.5) | 94.6 (92.1, 96.5) | 93.2 (91.8, 94.4) |
| April | 97.7 (96.8, 98.4) | 98.6 (97.3, 99.3) | 94.2 (91.8, 96.0) | 94.5 (93.3, 95.4) |
| **Moderna** | **98.7 (98.3, 99.0)** | **98.5 (97.7, 99.1)** | **97.3 (96.3, 98.1)** | **96.1 (95.5, 96.7)** |
| January/February | 98.6 (97.3, 99.4) | 98.6 (95.8, 99.7) | 94.7 (90.4, 97.4) | 95.4 (93.5, 96.8) |
| March | 98.6 (97.9, 99.1) | 99.1 (97.9, 99.7) | 97.3 (95.6, 98.4) | 95.9 (94.9, 96.7) |
| April | 98.8 (98.2, 99.3) | 98.0 (96.5, 98.9) | 98.3 (96.9, 99.1) | 96.6 (95.7, 97.3) |
| **Janssen** | **91.8 (88.6, 94.2)** | **91.1 (85.1, 95.0)** | **90.9 (84.9, 95.0)** | **89.3 (86.2, 91.8)** |
| March | 93.4 (88.7, 96.5) | 93.0 (83.7, 97.7) | 90.1 (79.5, 96.0) | 88.5 (83.4, 92.3) |
| April | 90.5 (85.9, 93.9) | 89.5 (80.6, 95.0) | 91.5 (83.1, 96.4) | 89.9 (85.8, 93.0) |

1. The restriction applied in this scenario reduced the number of events observed. Estimates of 100.0% reflect strata in which 0 hospitalizations among fully-vaccinated persons and should be interpreted both with caution and in conjunction with the lower-bound of the exact 95% confidence interval.

Table S7: Estimated vaccine effectiveness for laboratory-confirmed COVID-19 hospitalizations, using entire time-defined cohorts, without 90-day prior diagnosis exclusion applied (sensitivity analysis #4)

| ***Vaccine cohort*** | **May 2021** *% VE (95% CI)* | **June 2021** % VE (95% CI) | **July 2021** % VE (95% CI) | **August 2021** % VE (95% CI) |
|---|---|---|---|---|
| **18-49 years** | | | | |
| **Pfizer-BioNTech** | **96.1 (94.1, 97.6)** | **94.3 (90.2, 97.0)** | **95.6 (92.6, 97.6)** | **95.2 (93.6, 96.5)** |
| January/February | 97.8 (93.5, 99.5) | 98.2 (89.7, 100.0) | 96.0 (88.3, 99.2) | 92.6 (88.1, 95.7) |
| March | 98.9 (93.9, 100.0) | 97.3 (84.8, 99.9) | 96.1 (85.8, 99.5) | 94.9 (89.8, 97.8) |
| April | 94.8 (91.7, 96.9) | 92.0 (85.6, 96.0) | 95.3 (91.0, 97.9) | 96.4 (94.5, 97.8) |
| **Moderna** | **96.6 (94.3, 98.1)** | **91.4 (85.7, 95.2)** | **95.9 (92.4, 98.0)** | **97.3 (95.8, 98.4)** |
| January/February | 96.6 (92.0, 98.9) | 98.3 (90.5, 100.0) | 98.8 (93.2, 100.0) | 97.2 (94.2, 98.9) |
| March | 98.1 (93.1, 99.8) | 90.5 (75.4, 97.4) | 91.4 (79.9, 97.2) | 97.7 (94.2, 99.4) |
| April | 95.7 (91.4, 98.1) | 86.5 (74.9, 93.6) | 96.1 (90.0, 98.9) | 97.1 (94.5, 98.7) |
| **Janssen** | **95.7 (91.1, 98.3)** | **87.7 (75.6, 94.7)** | **94.5 (87.0, 98.2)** | **93.5 (89.7, 96.1)** |
| March | 90.2 (71.3, 98.0) | 51.0 (-7.4, 82.1) | 94.1 (67.1, 99.9) | 92.3 (80.2, 97.9) |
| April | 97.0 (92.2, 99.2) | 96.2 (86.2, 99.5) | 94.5 (86.0, 98.5) | 93.7 (89.5, 96.6) |
| **50-64 years** | | | | |
| **Pfizer-BioNTech** | **95.5 (94.1, 96.7)** | **94.8 (92.1, 96.8)** | **93.4 (90.7, 95.5)** | **94.7 (93.7, 95.7)** |
| January/February | 92.6 (87.7, 95.9) | 92.9 (83.2, 97.7) | 96.6 (89.9, 99.3) | 94.2 (91.1, 96.4) |
| March | 93.6 (88.9, 96.6) | 95.7 (87.4, 99.1) | 90.9 (81.8, 96.1) | 95.6 (92.8, 97.5) |
| April | 96.7 (95.2, 97.9) | 95.1 (91.7, 97.4) | 93.3 (89.9, 95.8) | 94.7 (93.3, 95.8) |
| **Moderna** | **97.1 (95.8, 98.1)** | **96.8 (94.1, 98.5)** | **96.4 (93.9, 98.1)** | **96.7 (95.6, 97.5)** |
| January/February | 98.2 (95.4, 99.5) | 100.0 (95.2, 100.0) | 97.9 (92.4, 99.7) | 96.7 (94.4, 98.3) |
| March | 100.0 (97.9, 100.0) | 98.4 (91.0, 100.0) | 97.4 (90.7, 99.7) | 97.8 (95.5, 99.1) |
| April | 95.6 (93.3, 97.3) | 94.8 (90.1, 97.7) | 95.4 (91.5, 97.8) | 96.2 (94.7, 97.4) |
| **Janssen** | **87.4 (82.2, 91.3)** | **90.4 (81.5, 95.6)** | **89.7 (81.9, 94.7)** | **92.4 (89.4, 94.6)** |
| March | 90.1 (80.4, 95.7) | 92.8 (74.0, 99.1) | 85.7 (66.3, 95.4) | 91.7 (85.5, 95.7) |
| April | 86.2 (79.7, 91.1) | 89.3 (77.7, 95.7) | 91.5 (82.2, 96.6) | 92.6 (89.1, 95.2) |
| **≥65 years** | | | | |
| **Pfizer-BioNTech** | **94.6 (93.8, 95.4)** | **93.4 (91.7, 94.8)** | **88.8 (86.5, 90.7)** | **88.4 (87.3, 89.5)** |
| January/February | 92.7 (90.4, 94.5) | 92.3 (88.1, 95.3) | 88.5 (83.5, 92.3) | 85.2 (82.5, 87.6) |
| March | 95.5 (94.1, 96.6) | 93.6 (90.8, 95.8) | 88.2 (84.4, 91.2) | 88.8 (87.0, 90.5) |
| April | 94.8 (93.6, 95.9) | 93.7 (91.2, 95.6) | 89.4 (86.2, 92.0) | 89.6 (88.0, 91.0) |
| **Moderna** | **97.0 (96.4, 97.6)** | **96.2 (94.9, 97.2)** | **94.8 (93.4, 96.0)** | **93.6 (92.8, 94.4)** |
| January/February | 97.2 (95.4, 98.4) | 95.9 (91.8, 98.2) | 91.1 (85.7, 94.9) | 92.7 (90.3, 94.6) |
| March | 97.3 (96.3, 98.0) | 97.3 (95.5, 98.5) | 95.5 (93.3, 97.1) | 93.7 (92.4, 94.7) |
| April | 96.7 (95.7, 97.5) | 95.1 (92.9, 96.8) | 95.4 (93.2, 97.0) | 93.8 (92.6, 94.9) |
| **Janssen** | **84.5 (80.2, 88.1)** | **79.6 (70.9, 86.2)** | **81.3 (72.8, 87.6)** | **81.7 (77.6, 85.2)** |
| March | 89.7 (83.9, 93.8) | 79.2 (64.7, 88.7) | 81.9 (68.0, 90.7) | 81.8 (75.3, 86.8) |
| April | 80.6 (74.2, 85.8) | 80.0 (68.1, 88.2) | 80.8 (69.0, 88.9) | 81.6 (76.1, 86.1) |
